# Combining faecal immunochemical testing with blood test results to identify patients with symptoms at risk of colorectal cancer: a consecutive cohort of 16,604 patients tested in primary care

**DOI:** 10.1101/2021.10.22.21263919

**Authors:** Diana R Withrow, Brian Shine, Jason Oke, Andres Tamm, Tim James, Eva Morris, Jim Davies, Steve Harris, James E East, Brian D Nicholson

**Affiliations:** Nuffield Department of Primary Care Health Sciences, Medical Sciences Division, University of Oxford, Radcliffe Observatory Quarter, Oxford, OX2 6GG, United Kingdom; Department of Clinical Biochemistry, John Radcliffe Hospital, Oxford University Hospitals NHS Foundation, Oxford, United Kingdom; Nuffield Department of Population Health, Big Data Institute, University of Oxford, Oxford, United Kingdom; Department of Computer Science, Big Data Institute, University of Oxford, Oxford, United Kingdom; Oxford BRC Informatics Theme, Big Data Institute, University of Oxford, Oxford, United Kingdom; Translational Gastroenterology Unit, John Radcliffe Hospital, University of Oxford, Oxford, United Kingdom

## Abstract

**Objective:** Faecal immunochemical tests (FITs) are used to triage primary care patients with low risk colorectal cancer symptoms for referral to colonoscopy. The aim of this study was to determine whether combining FIT with routine blood test results could improve the performance of FIT in the primary care setting.

**Design:** Results of all consecutive FITs requested by primary care providers between March 2017 and December 2020 were retrieved from the Oxford University Hospitals NHS Foundation Trust. Demographic factors (age, sex), reason for referral, and results of blood tests within 90 days were also retrieved. Patients were followed up for incident colorectal cancer in linked hospital records. The sensitivity, specificity, positive and negative predictive values of FIT alone, FIT paired with blood test results, and several multivariable FIT models, were compared.

**Results:** Among 16,604 eligible patients, 139 colorectal cancers were diagnosed (0.8%). Sensitivity and specificity of FIT alone at a threshold of 10 µg Hb/g were 92.1% and 91.5% respectively. Compared to FIT alone, blood test results did not improve the performance of FIT. Pairing blood test abnormalities with FIT reduced the number of abnormal results needed to detect one cancer but increased the number of cancers missed. Multivariable models retaining FIT, sex, and mean cell volume performed similarly to FIT alone.

**Conclusion:** FIT is a highly sensitive tool for identifying higher risk individuals presenting to primary care with lower risk symptoms. Combining blood test results with FIT does not appear to lead to better discrimination for colorectal cancer than using FIT alone.

## Introduction

Diagnosing colorectal cancer in patients who present to primary care can be challenging because many of the symptoms of colorectal cancer are shared with other, less serious causes. Colonoscopy is the definitive test to diagnose colorectal cancer, but referring all patients with symptoms of possible colorectal cancer for colonoscopy would cause significant strain on health care resources and present unnecessary risks to patients^1^. In 2017, the faecal immunochemical test (FIT) was recommended by the National Institute for Health and Care Excellence (NICE) as a triage test for patients presenting to primary care with low risk symptoms of possible colorectal cancer ^2^. The evidence underpinning that recommendation was drawn primarily from higher risk populations and there was limited evidence about how it would perform in primary care ^3-5^.

There has been a rapid increase in publications about FIT use in symptomatic patients over the last five years^3 6 7^. FIT has consistently been shown to have high sensitivity and specificity for colorectal cancer at a threshold of 10µg Hb/g faeces or lower, in primary and secondary care^7-9^. Despite a high negative predictive value, nearly one in ten colorectal cancers will be missed using FIT alone to select who should be referred for investigation ^10^. Developing strategies to identify symptomatic people with FIT negative colorectal cancer has become an urgent priority due to the increased use of FIT to defer or decline colorectal investigation during the COVID-19 pandemic^11^. Furthermore, as the number of colorectal cancer presentations is expected to increase, and health care resources continue to be strained by ongoing effects of the pandemic, efforts to reduce unnecessary referrals by increasing specificity would be especially worthwhile^11^.

Clinical prediction models are one strategy to achieve these aims. However, the faecal haemoglobin age and sex test (FAST) score did not improve utility over FIT alone^12^. FIT has also been shown to outperform multivariable models including age, sex and symptoms prompting urgent cancer referral^13^. Combining commonly used blood tests with FIT could further optimise the triage of symptomatic patients in primary care for colorectal cancer investigation ^14 15^.

Using the largest existing UK cohort of symptomatic patients tested with FIT in primary care, the aim of this study was to assess whether complementing FIT with blood test values could improve the predictive performance of FIT^16^.

## Methods

### Study design

#### Population/Setting

Data were retrieved from Oxford University Hospitals NHS Foundation Trust (OUH). OUH serves all primary care clinicians in the county of Oxfordshire, UK, with a population of approximately 660,000. Based at the John Radcliffe Hospital, the Clinical Biochemistry Laboratory performs over 8 million tests a year. This study was registered as a service evaluation on the OUH Datix register (CSS-BIO-3 4730).

#### FITs

All consecutive FIT results (measured in µg Hb/g faeces) between March 2017 and December 21^st^ 2020 were retrieved retrospectively from the OUH Clinical Biochemistry Laboratory Information Management System. The end date for eligible FITs was determined to allow a minimum of 6 months follow-up for all participants.

After restricting to FITs requested by primary care clinicians and the first FIT in any given individual, FITs were retained for inclusion in this analysis if the five most common “core” blood tests (haemoglobin, platelets, white cell count, MCH and MCV) were available, patients were aged 18 or older, had known sex, and had non-missing FIT results (Figure 1).

**Figure 1.**
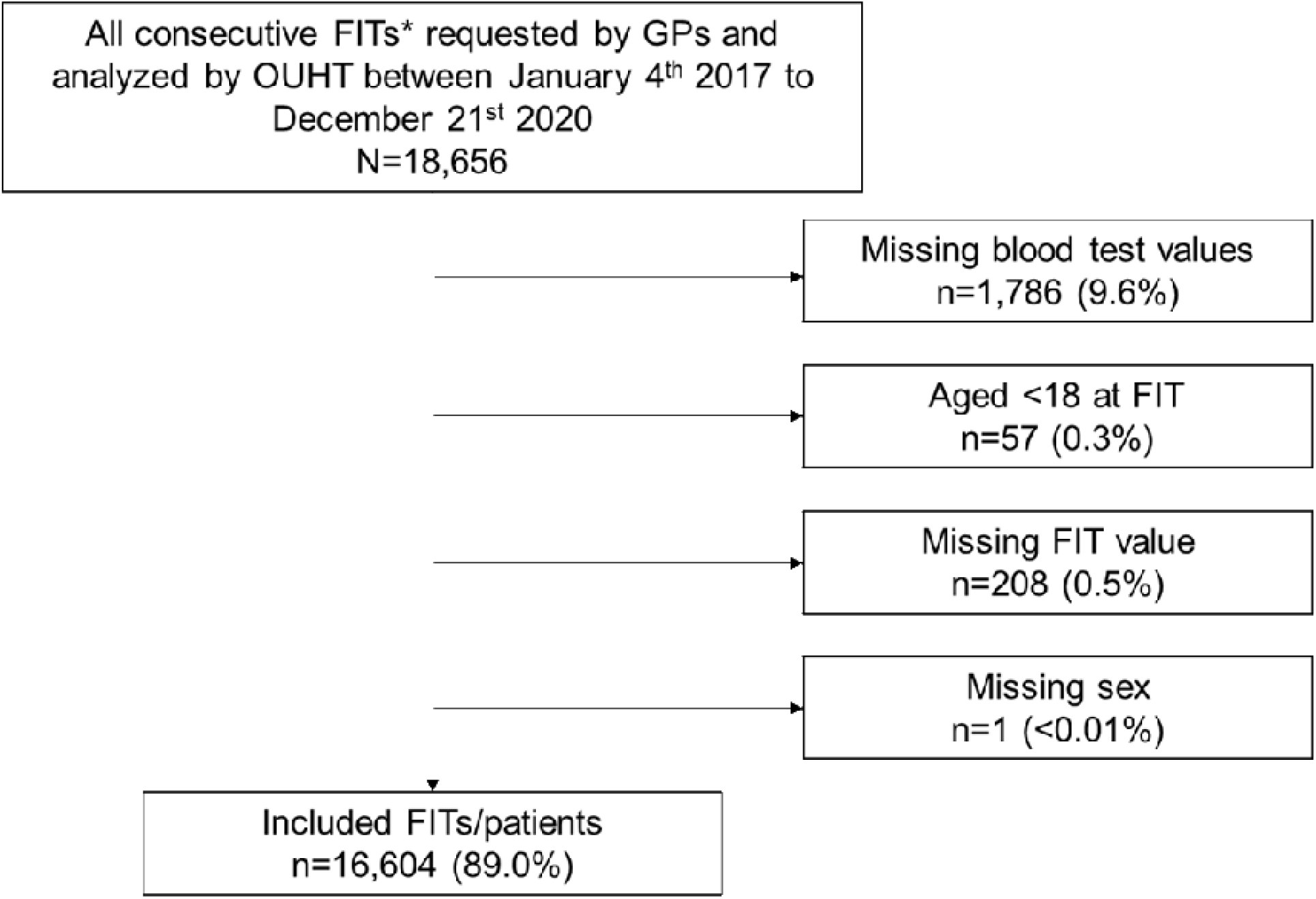
Selection criteria for inclusion. *First FITs per individual.

.Faecal specimens were collected into standard pots by patients in primary care and referred to the central laboratory where sampling was undertaken using the Extel Hemo-Auto MC device. Prepared samples were analysed for FIT using the HM-JACKarc analyser (Hitachi Chemical Diagnostics Systems Co., Ltd, Tokyo, Japan and distributed in the UK by Alpha Labs Ltd, Eastleigh, Hants) a method recommended for use by NICE^2^. The method had a calibration range of 7-400 µg Hb/g faeces. Immunoassay reproducibility assessed across 12 months had a coefficient of variation (CV) of between 4.4 and 8.8%. The overall imprecision of the process including sampling variation was between 7.0 and 13.5 CV% ^17^ FIT samples were assayed and recorded prior to and independent of the any subsequent pathology findings.

#### Additional variables

Age, sex, clinical indication, and results of contemporaneous blood tests were retrieved for each patient. To extract the clinical indication, free text fields for the reason for referral were searched for common indications (abdominal pain, bloating, blood in stool, change in bowel habit, constipation, diarrhoea, family history of cancer, fatigue, melaena, rectal pain, and weight loss) using numerous permutations of spelling and phrasing. A 20% sample was reviewed to ensure validity of free text coding.

Blood test results reported less than 60 days prior to or 30 days post FIT were retrieved. The most routinely used blood tests and those with a hypothesized relationship with colorectal cancer risk were selected for analysis (haemoglobin, platelets, white cell count, mean cell haemoglobin (MCH), mean cell volume (MCV); serum ferritin and c-reactive protein [CRP])^18^. The same analytical methods for the blood tests were used throughout the study period: full blood count, including haemoglobin, platelets, white cell count, MCH and MCV were analysed using a Sysmex XN analyser (Sysmex UK Ltd, Milton Keynes, UK); ferritin using an Abbott Architect i2000 and CRP using the Abbott Architect c16000 (both Abbott Diagnostics UK, Maidenhead, UK).

#### Outcomes

A composite reference standard was used as not all patients tested with FIT in primary care are referred for definitive testing. A reliance on definitive testing alone would lead to verification bias for FIT positive patients. The composite reference standard incorporated the review of multiple linked databases (hospital clinical records, pathology results, and endoscopy and radiology reports) for evidence of a new colorectal cancer diagnosis within the follow-up period, which was 6 months in the primary analysis. Database review was independent of FIT value.

#### Patient & Public Involvement

No patients were directly involved in designing the research question or in conducting the research. A patient advocate provided feedback on interpretation of the results and key messages. Our findings will be disseminated to patients and the public through the NIHR BRC, Nuffield Department of Primary Care Health Sciences, Oxford Cancer, and OxCODE.

### Statistical Analysis

Three approaches were investigated to optimise FIT.

1. FIT alone - dichotomous FIT at a cut-off of greater than or equal to 2 or 10 µg Hb/g faeces;
2. FIT-Blood test pairs – dichotomous FIT and dichotomous blood test result; A test was considered positive if patients fell above the cut-off value for FIT (2 or 10 µg Hb/g faeces) and had an abnormal blood test result. The threshold for abnormal blood tests were pre-specified based on standard clinical practice^19^.
3. Multivariable FIT – modeling including FIT, blood tests, age and sex.

Logistic regression was used to generate predicted probabilities of colorectal cancer. Backward stepwise selection was used to select covariates. Because serum ferritin and CRP were only available for a subset of cases, stepwise selection was conducted on an imputed dataset with 10 replicates using predictive mean matching. In models where CRP or serum ferritin were retained, coefficients for each variable in the imputed and complete case datasets were compared and if similar, the model results from the complete case dataset were reported. The three modeling approaches are defined below.

Model A: FIT, age, and blood test results (continuous); sex (dichotomous).

Model B: FIT and blood test results (dichotomous); age (categorical); and sex (dichotomous)

Model C: FIT (spline), age (continuous), sex and blood tests (dichotomized).

The restricted cubic spline function for FIT was specified to have knots at 2, 10, 50 and 100 µg Hb/g faeces. Four knots were selected to yield a model with at least 20 events per variable, to minimize optimism bias^20^. Ninety-five percent confidence intervals were estimated using the Wilson Score method^21^. The positive predictive value (PPV) and negative predictive value (NPV) were additionally expressed as the number of positive FITs to detect one cancer (number needed to scope) and the cancer miss rate per 10,000 negative tests. To permit a comparison of model performance, the probability cut-off to determine a positive result was selected to match the sensitivity of the FIT alone at a cut-off of 10 µg Hb/g faeces.

### Sensitivity & Subgroup Analyses

The FIT alone approach was applied to subgroups defined by FIT date (prior to or during the COVID-19 pandemic), age-group (<40, >50, >60, >70, >80), sex, blood test results and clinical indication. Each of the approaches 1, 2, and 3 outlined above were replicated with 12 months of follow-up.

## Results

### Descriptive

A total of 16,604 of 18,656 available FITs (89%) were included in the study. Included patients were representative of the overall sample (Table 1). Study subjects had a median age of 61 and were 58% female. One-hundred and thirty-nine (139) cancers were diagnosed within six months of the FIT test (0.8%). Patients who were diagnosed with cancer were older (median age 72) and more likely to be male (60%), to have a FIT ≥10 µg Hb/g faeces and/or to have abnormal blood tests (Table 1, Figure 2, Supplementary Table 1).

**Table 1.** Characteristics of patients receiving symptomatic FIT tests by study inclusion status and outcome.

|  | All FIT tests |  | Included |  | Cancer |  |
| --- | --- | --- | --- | --- | --- | --- |
|  | n | % | n | % | n | % |
| **Total** | 18,656 | 100 | 16,604 | 100 | 139 | 100 |
| **Age** |  |  |  |  |  |  |
| 0-18 | 95 | 1 | 0 | 0 | 0 | 0 |
| 18-39.9 | 1,651 | 9 | 1,390 | 8 | 9 | 6 |
| 40-49.9 | 2,553 | 14 | 2,278 | 14 | 12 | 9 |
| 50-59.9 | 4,679 | 25 | 4,181 | 25 | 20 | 14 |
| 60-69.9 | 3,186 | 17 | 2,892 | 17 | 21 | 15 |
| 70-79.9 | 3,711 | 20 | 3,330 | 20 | 36 | 26 |
| ≥ 80 | 2,781 | 15 | 2,533 | 15 | 41 | 29 |
| Median (IQR) | 61 (50, 74) |  | 61 (51, 75) |  | 72 (57, 81) |  |
| **Sex** |  |  |  |  |  |  |
| Male | 7,926 | 42 | 7,019 | 42 | 83 | 60 |
| Female | 10,728 | 58 | 9,585 | 58 | 56 | 40 |
| Unknown | 2 | 0 | 0 | 0 | 0 | 0 |
| **FIT (µg Hb/g)** |  |  |  |  |  |  |
| 0-1.9 | 15,298 | 82 | 13,757 | 83 | 5 | 4 |
| 2-9.9 | 1,409 | 8 | 1,318 | 8 | 6 | 4 |
| 10-99.9 | 1,072 | 6 | 1,023 | 6 | 51 | 37 |
| ≥ 100 | 539 | 3 | 506 | 3 | 77 | 55 |
| Missing | 338 | 2 | 0 | 0 |  |  |
| Median (IQR) | 0 (0, 0.7) |  | 0.2 (0, 0.8) |  | 135.5 (33.4, 450) |  |
| **Blood test results\*** |  |  |  |  |  |  |
| Low haemoglobin<sup>a</sup> | 5,186 | 31 | 5,076 | 31 | 72 | 52 |
| High platelets<sup>b</sup> | 556 | 3 | 546 | 3 | 13 | 9 |
| High white cells<sup>c</sup> | 832 | 5 | 820 | 5 | 9 | 6 |
| Low mean cell haemoglobin<sup>d</sup> | 2,792 | 17 | 2,730 | 16 | 47 | 34 |
| Low mean cell volume<sup>e</sup> | 1,014 | 6 | 980 | 6 | 30 | 22 |
| Any abnormal FBC | 6,521 | 35 | 6,392 | 38 | 81 | 58 |
| Low serum ferritin<sup>f</sup> | 2,015 | 22 | 1,962 | 22 | 36 | 40 |
| High serum ferritin<sup>g</sup> | 457 | 5 | 444 | 5 | 3 | 3 |
| High C-reactive protein<sup>h</sup> | 1,748 | 14 | 1,720 | 14 | 31 | 31 |
| **Symptoms - GP reported** |  |  |  |  |  |  |
| Abdominal pain | 3,299 | 18 | 2,941 | 18 | 23 | 17 |
| Blood in stool | 1,759 | 9 | 1,451 | 9 | 22 | 16 |
| Melaena | 298 | 2 | 238 | 1 | 0 | 0 |
| Change in bowel habit | 7,511 | 40 | 6,656 | 40 | 45 | 32 |
| Diarrhoea | 2,651 | 14 | 2,315 | 14 | 13 | 9 |
| Constipation | 722 | 4 | 608 | 4 | 1 | 1 |
| Fatigue | 199 | 1 | 193 | 1 | 1 | 1 |
| Rectal pain | 106 | 1 | 95 | 1 | 0 | 0 |
| Bloating | 594 | 3 | 541 | 3 | 2 | 1 |
| Family history of cancer | 342 | 2 | 263 | 2 | 2 | 1 |
| Weight loss | 1,448 | 8 | 1,348 | 8 | 9 | 6 |
| **Blood - GP reported** |  |  |  |  |  |  |
| Anaemia (any) | 4,517 | 24 | 4,272 | 26 | 48 | 35 |
| Iron deficiency anaemia | 1,926 | 10 | 1,793 | 11 | 18 | 13 |
| Thrombocytosis | 216 | 1 | 204 | 1 | 2 | 1 |

IQR: Interquartile range. \* percent with non-missing values <sup>a</sup> <130 g/L in men and <120 g/L in women; <sup>b</sup> >400 µL/L; <sup>c</sup> <11,000/mL; <sup>d</sup> <27.4 pg/cell; <sup>e</sup> <80 fL; <sup>f</sup> <20 ng/mL; <sup>g</sup> ≥350 ng/mL; <sup>h</sup> >10
Any abnormal full blood count (FBC) refers to any abnormal result of heamoglobin, platelets, white cells, mean cell haemoglobin and mean cell volume.
**Note:** Serum ferritin and c-reactive protein tests were only conducted for a subset of patients (n=8,922 and 12,201 respectively)

**Figure 2.**
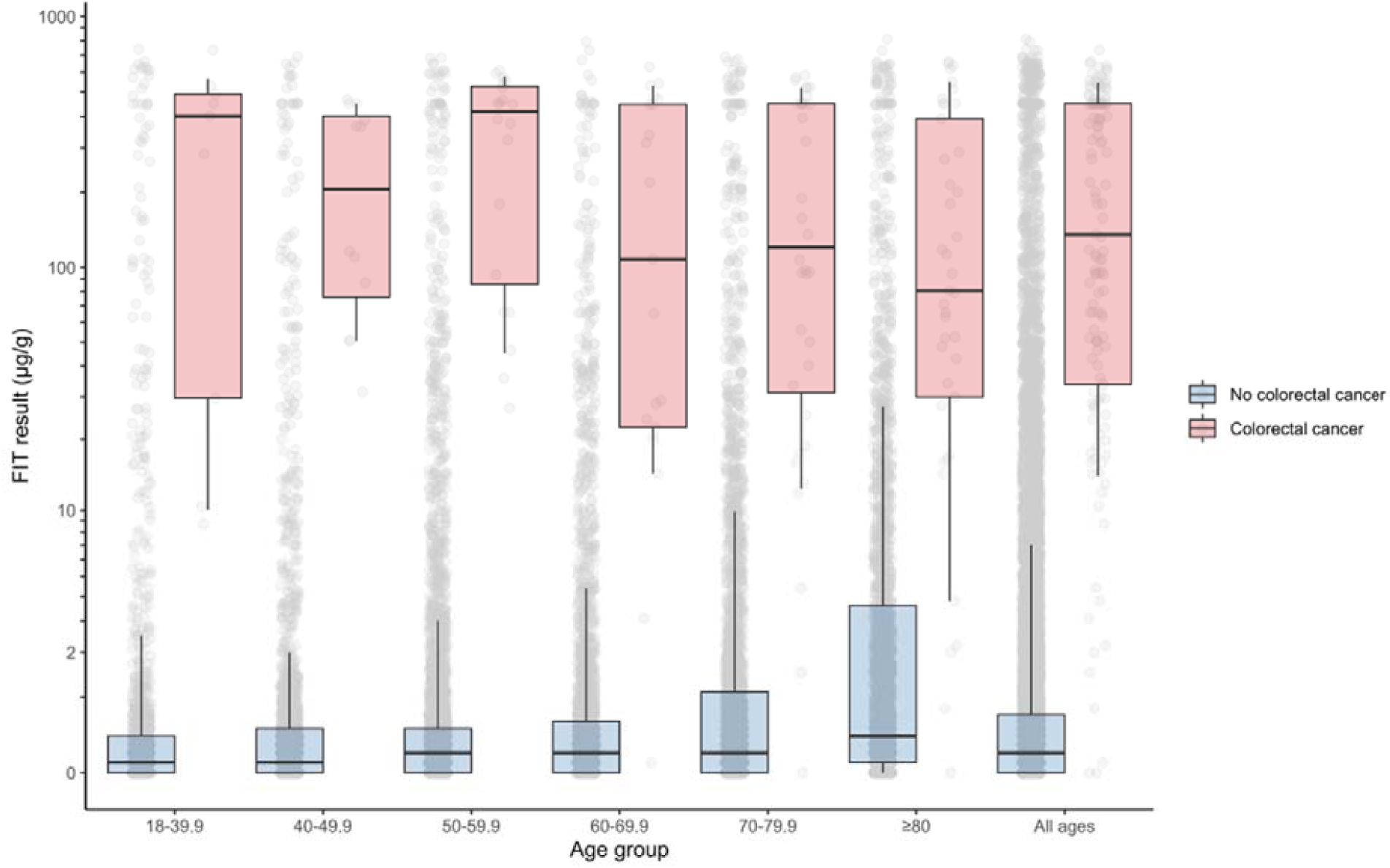
Distribution of FIT score by age and outcome. Boxes indicate median and interquartile range. Whiskers indicate 10th and 90th percentiles. The shape of the distribution corresponds to log10(FIT + 1) whereas tick marks are drawn at actual FIT values.

Ninety percent of patients had at least one reported clinical indication, most commonly change in bowel habit (40%) then anaemia (26%) and abdominal pain (18%, Table 1). The most common clinical indications in people with cancer were anaemia (35%), change in bowel habit (32%), blood in stool (16%) and abdominal pain (17%).

Low haemoglobin was the most common abnormal blood test result (31% of all patients, 52% of those with a subsequent cancer diagnosis, Table 1) followed by low MCH (16% and 34% respectively).

#### 1) FIT alone

At a FIT threshold of 2 µg Hb/g faeces sensitivity was 96.4% (95% CI: 91.9-98.5), specificity 83.5% (95% CI: 82.9-84.1), PPV 4.7% (95% CI: 4.0-5.5), and NPV 100% (95% CI: 99.9-100) (Table 2, Supplementary Table 2). One cancer was detected for every twenty-one positive FITs, and the cancer miss rate was 4 cancers per 10,000 negative tests (Table 2).

**Table 2.** Test performance as measured by positive and negative predictive value (PPV, NPV), sensitivity, specificity, positive FITs per cancer detected and cancer miss rate per 10,000 negative tests. FIT alone and threshold-based approach to FIT-blood test pairs.

| Test criteria |  | PPV (95% CI) | NPV (95% CI) | Sensitivity (95% CI) | Specificity (95% CI) | Positive FITs to detect one cancer | Cancer miss rate per 10,000 negative tests |
| --- | --- | --- | --- | --- | --- | --- | --- |
| FIT alone |  |  |  |  |  |  |  |
| FIT≥2 μg Hb/g | | 4.7% (4, 5.5) | 100.0% (99.9, 100) | 96.4% (91.9, 98.5) | 83.5% (82.9, 84.1) | 21 | 4 |
| FIT≥10 μg Hb/g | | 8.4% (7.1, 9.9) | 99.9% (99.9, 100) | 92.1% (86.4, 95.5) | 91.5% (91.1, 91.9) | 12 | 7 |
| FIT-Blood Test Pairs |  |  |  |  |  |  |  |
| FIT≥2 μg Hb/g AND | Low haemoglobin<sup>a</sup> | 5.1% (4.3, 6.2) | 99.9% (99.8, 99.9) | 89% (82.2, 93.5) | 83.4% (82.7, 84) | 17 | 44 |
|  | High platelets<sup>b</sup> | 5.8% (4.6, 7.3) | 99.6% (99.4, 99.7) | 51% (42.9, 59.2) | 93.0% (92.6, 93.4) | 11 | 77 |
|  | High white cells<sup>c</sup> | 8.7% (5.2, 14.4) | 92.8% (91.5, 93.9) | 9.4% (5.5, 15.3) | 92.3% (91, 93.4) | 29 | 80 |
|  | Low mean cell haemoglobin<sup>d</sup> | 3.4% (1.7, 6.6) | 99.2% (99.1, 99.3) | 5.8% (2.9, 10.9) | 98.6% (98.4, 98.8) | 12 | 57 |
|  | Low mean cell volume<sup>e</sup> | 8.0% (6.1, 10.5) | 99.4% (99.3, 99.5) | 33.8% (26.5, 42) | 96.7% (96.4, 97) | 6 | 66 |
|  | Any abnormal FBC | 15.9% (11.4, 21.8) | 99.3% (99.2, 99.4) | 21.6% (15.6, 29.1) | 99.0% (98.9, 99.2) | 19 | 40 |
|  | Low serum ferritin<sup>f</sup> | 5.4% (4.3, 6.7) | 99.6% (99.5, 99.7) | 56.8% (48.5, 64.8) | 91.6% (91.1, 92) | 10 | 63 |
|  | High serum ferritin<sup>g</sup> | 10.0% (7.3, 13.6) | 99.4% (99.2, 99.5) | 40.0% (30.5, 50.3) | 96.3% (95.9, 96.7) | 33 | 99 |
|  | High C-reactive protein<sup>h</sup> | 3.0% (1, 8.5) | 99.0% (98.8, 99.2) | 3.3% (1.1, 9.3) | 98.9% (98.7, 99.1) | 18 | 59 |
| FIT≥10 μg Hb/g AND | Low haemoglobin<sup>a</sup> | 5.6% (4, 7.8) | 99.4% (99.3, 99.5) | 31.0% (22.8, 40.6) | 95.7% (95.3, 96) | 10 | 44 |
|  | High platelets<sup>b</sup> | 10.3% (8.2, 12.8) | 99.6% (99.4, 99.7) | 49.6% (41.5, 57.8) | 96.4% (96.1, 96.6) | 8 | 77 |
|  | High white cells<sup>c</sup> | 12.2% (7.1, 20.2) | 99.2% (99.1, 99.4) | 8.6% (5, 14.5) | 99.5% (99.4, 99.6) | 17 | 80 |
|  | Low mean cell haemoglobin<sup>d</sup> | 5.8% (2.9, 10.9) | 99.2% (99.1, 99.3) | 5.8% (2.9, 10.9) | 99.2% (99.1, 99.3) | 7 | 58 |
|  | Low mean cell volume<sup>e</sup> | 13.5% (10.3, 17.6) | 99.4% (99.3, 99.5) | 32.4% (25.2, 40.5) | 98.3% (98, 98.4) | 4 | 67 |
|  | Any abnormal FBC | 23.3% (16.7, 31.7) | 99.3% (99.2, 99.4) | 20.1% (14.3, 27.6) | 99.4% (99.3, 99.5) | 10 | 39 |
|  | Low serum ferritin<sup>f</sup> | 9.6% (7.7, 11.8) | 99.6% (99.5, 99.7) | 55.4% (47.1, 63.4) | 95.6% (95.3, 95.9) | 6 | 62 |
|  | High serum ferritin<sup>g</sup> | 17.0% (12.5, 22.6) | 99.4% (99.2, 99.5) | 40.0% (30.5, 50.3) | 98.0% (97.7, 98.3) | 15 | 98 |
|  | High C-reactive protein<sup>h</sup> | 6.5% (2.2, 17.5) | 99.0% (98.8, 99.2) | 3.3% (1.1, 9.3) | 99.5% (99.3, 99.6) | 13 | 62 |

<sup>a</sup> <130 g/L in men and <120 g/L in women; <sup>b</sup> >400 $\mu$ L/L; <sup>c</sup> <11,000/mL; <sup>d</sup> <27.4 pg/cell; <sup>e</sup> <80 fL; <sup>f</sup> <20 ng/mL; <sup>g</sup> ≥ 350 ng/mL; <sup>h</sup> >10 mg/L. CI: Confidence interval.
Note: Serum ferritin and c-reactive protein tests were only conducted for a subset of patients (n=8,923 and 12,202 respectively)

At 10 µg Hb/g faeces, sensitivity was 92.1% (95% CI: 86.4-95.5), specificity 91.5% (95% CI: 91.1-91.9), PPV 8.4% (95%CI: 7.1-9.9) and NPV 99.9% (95% CI: 99.9-100) (Table 2, Supplementary Table 2). One cancer was detected for every twelve positive FITs, and a miss rate of 7 cancers per 10,000 negative tests (Table 2).

#### 2) FIT-Blood Test Pairs

Sensitivity ranged from 3.3% (FIT≥2 or 10 µg Hb/g faeces and raised CRP) to 56.8% (FIT≥2 µg Hb/g faeces and low serum ferritin) for pairings of FIT and blood tests. Specificity was higher for almost all pairings compared to a FIT-alone approach leading to fewer positives being needed to detect one cancer. However, the cancer miss rate per 10,000 tests increased 14-fold compared to a FIT alone approach (Table 2).

#### 3) Multivariable FIT

A. Model A (with continuous FIT): sex and continuous variables for age, serum ferritin, platelets and CRP were retained. Specificity was 45.9% (95% CI 44.7-47.1), compared to 90.0% for FIT alone (in the subset with serum ferritin and CRP), leading to one cancer in every 57 positive tests compared to one in 12 in the FIT-only approach (Table 3, Supplementary Table 2).
B. Model B (dichotomous FIT, blood tests): FIT, sex and low MCV were retained. Specificity was 90.1% (95% CI: 89.6-90.5), similar to FIT alone at FIT≥10 µg Hb/g faeces, leading to 14 positive tests to detect one cancer.
C. Model C (FIT spline): FIT, sex, and low MCV were retained. Specificity was 91.5% (95% CI: 91.1-91.9) with one cancer detected for every 12 positive FITs.

**Table 3.**
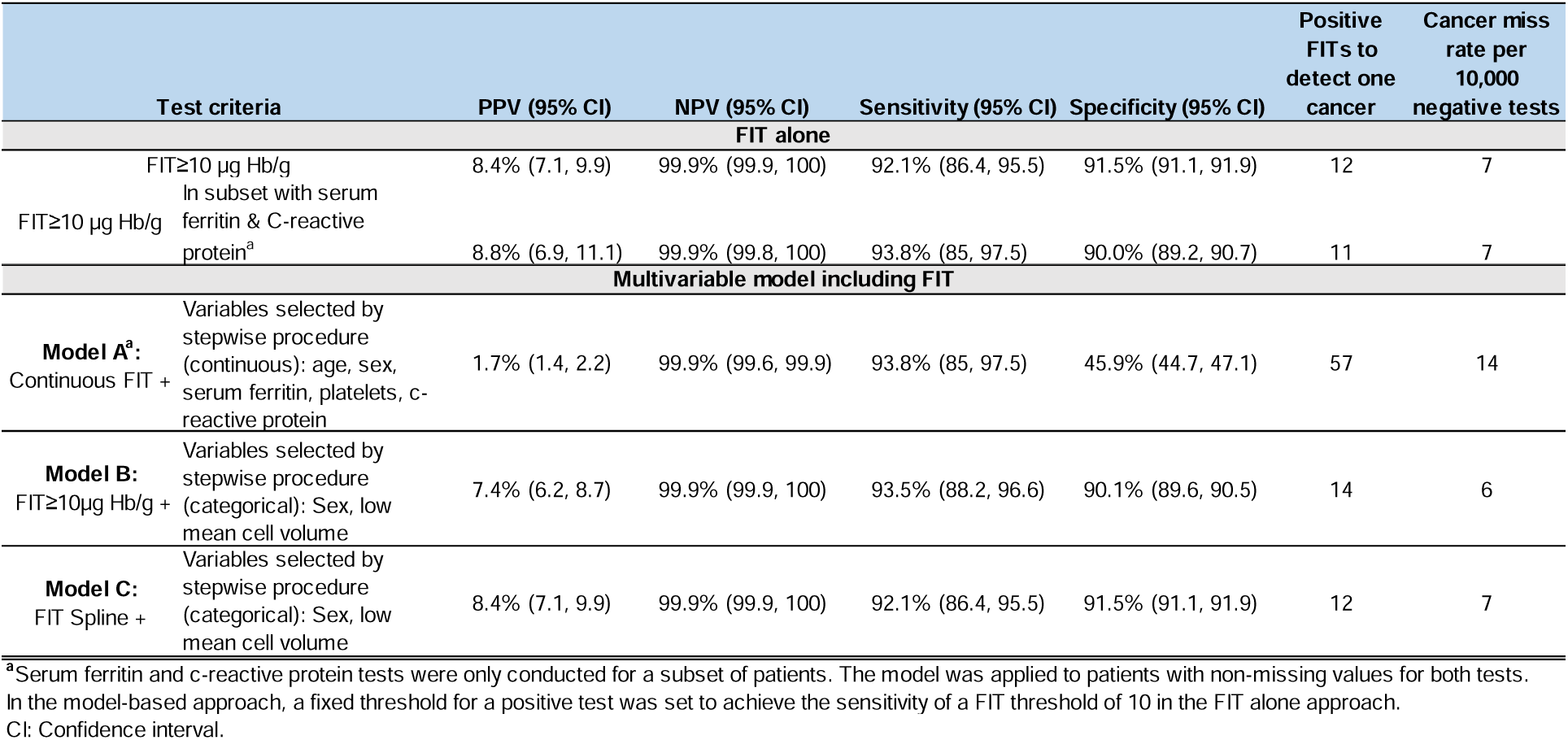
Test performance as measured by positive and negative predictive value (PPV, NPV), sensitivity, specificity, positive FITs per cancer detected and cancer miss rate per 10,000 negative tests. FIT alone and model-based approach.

**Table 4.** Clinical characteristics of patients who had a false-negative FIT at a threshold of 10 µg Hb/g faeces.

|  | 1 | 2 | 3 | 4 | 5 | 6 | 7 | 8 | 9 | 10 | 11 |
| --- | --- | --- | --- | --- | --- | --- | --- | --- | --- | --- | --- |
| **Age** | 18-40 | 80-89 | 80-89 | 60-69 | 70-79 | 60-69 | 70-79 | 70-79 | 80-89 | 80-89 | 80-89 |
| **Sex** | Male | Male | Female | Male | Male | Female | Female | Female | Male | Female | Male |
| **FIT result (µg Hb/g faeces)** | 8.7 | 3.8 | 2 | 0.1 | 0 | 3.1 | 4.4 | 1.5 | 2.2 | 0.8 | 0 |
| **Tumor location** | Splenic flexure | Sigmoid colon | Left descending colon | Rectum | Rectum | Unk | Unk | Rectum | Unk | Distal ascending colon | Sigmoid colon |
| **Interval from FIT to diagnosis (days)** | 27 | 25 | 36 | 55 | 21 | 7 | 20 | 92 | 75 | 24 | 29 |
| **Abnormal blood tests** |  |  |  |  |  |  |  |  |  |  |  |
| High C-reactive protein | Yes | Yes | Yes | No | No | Yes | Yes | No | No | No | Unk |
| Low haemoglobin | Yes | No | No | No | No | No | No | No | Yes | No | Yes |
| Low mean cell haemoglobin | Yes | No | No | No | No | No | No | No | Yes | No | No |
| Low mean cell volume | Yes | No | No | No | No | No | No | No | Yes | No | No |
| High platelets | Yes | No | No | No | No | No | No | No | No | No | No |
| High white cells | No | No | No | No | Yes | No | No | No | No | No | No |
| Low serum ferritin | No | Unk | No | Unk | Unk | Unk | No | Unk | No | Unk | No |
| High serum ferritin | No | Unk | No | Unk | Unk | Unk | No | Unk | No | Unk | No |
| **GP-reported symptoms** |  |  |  |  |  |  |  |  |  |  |  |
| Change in bowel habit | No | No | No | Yes | Yes | Yes | Yes | Yes | No | Yes | No |
| Diarrhoea | No | No | No | No | Yes | No | No | No | No | No | No |
| Constipation | No | No | No | No | No | No | No | No | No | No | No |
| Anaemia | Yes | No | No | No | Yes | No | No | No | No | No | Yes |
| Weight loss | Yes | No | No | No | Yes | No | No | No | Yes | No | No |
| Abdominal pain | No | Yes | No | No | No | No | No | No | No | No | No |
| Blood in stool | No | No | No | No | No | No | No | No | No | No | No |
| Abdominal mass | No | No | No | No | No | No | No | No | No | No | No |
| Melaena | No | No | No | No | No | No | No | No | No | No | No |
| Thrombocytosis | No | No | No | No | No | No | No | No | No | No | No |
| Fatigue | No | No | No | No | No | No | No | No | No | No | No |
| Bloating | No | No | No | No | No | No | No | No | No | No | No |
| Family history | No | No | No | No | No | No | No | No | No | No | No |

Note: Serum ferritin and c-reactive protein were only assessed in a subset of patients. Unk: Unknown.

In summary, Models B and C performed similarly to FIT alone but no approach that integrated blood test results improved the overall performance of FIT. Odds ratios for the predictors and the log likelihood and Area Under the Curve for each model are provided in Supplementary Table 3. A plot of apparent calibration did not reveal any causes for concern.

The age-specific probabilities of colorectal cancer by sex and FIT score based on Model C are illustrated in Figure 3. For males and females, the probability of colorectal cancer reached 3% (the cut-off specified to prompt urgent investigation by NICE^22^) at FIT values of 17 and 25 respectively. There were no significant differences by age since age was not a significant predictor of cancer risk after accounting for FIT (Supplementary Table 3).

**Figure 3.**
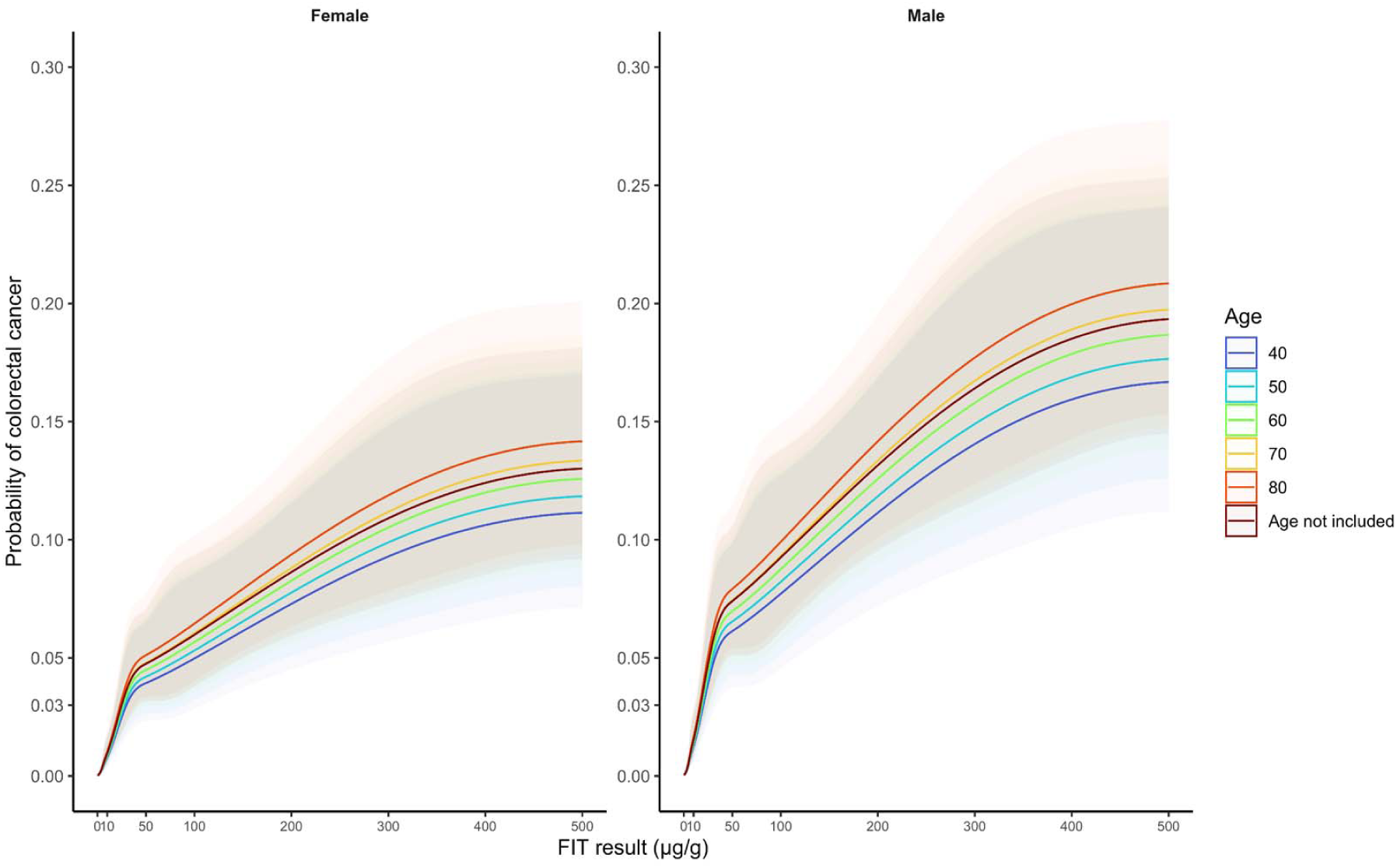
Probability of colorectal cancer by sex, age, and FIT score with 95% confidence intervals indicated with shading (See Model C). The restricted cubic spline function was specified to have knots at FIT values of 2, 10, 50 and 100.

### FIT-negative cancers

The characteristics of the 11 patients with false negative tests at a FIT threshold of 10 µg Hb/g faeces are provided in Table 3. Ten had at least one GP-reported clinical indication with the most common being change in bowel habit (n=6). Eight of the 11 had at least one abnormal blood test with the most common being raised CRP (5 of 10 with known values). Median days from FIT to cancer diagnosis was 27 days among false negatives (interquartile range 21 to 55) compared to 34 (21, 64) among persons diagnosed with cancer overall.

### Subgroup and sensitivity analyses

Patient demographics, clinical indication, prevalence of abnormal blood tests, FIT score, and performance of FIT were largely consistent prior to and during the COVID-19 pandemic (Supplementary Tables 4, 5). The median age of persons undergoing FIT was older during COVID (64 vs. 59 years) but the interquartile range was similar (51 to 76 vs. 51 to 74). There were no significant differences in sensitivity, specificity, PPV or NPV of FIT.

PPV was higher among males than females, but the confidence intervals for the two sexes overlapped at a threshold of 10 µg Hb/g faeces. At 2 µg Hb/g faeces, PPV and NPV decreased with increasing age. At 10 µg Hb/g faeces, PPV and NPV were largely consistent by age group (Supplementary Table 6).

There was no evidence that the PPV of FIT was significantly higher within subgroups defined by clinical indication or blood test other than MCV (Supplementary Table 6).

Results did not meaningfully change when the follow-up period was extended to 12 months (Supplementary Tables 7 and 8).

## Discussion

### Summary of findings

In this large cohort of symptomatic patients tested with FIT in primary care, neither age, nor blood test results remained strong enough predictors of colorectal cancer to improve on the performance of FIT. While the number of false positives could be reduced by taking into account blood tests, the large associated increase in false negatives outweighed the benefit. In addition, there was no evidence to suggest that using clinical indication as a rule-out or rule-in factor would improve the efficiency of FIT triage.

It may seem counter-intuitive that blood tests known to be associated with colorectal cancer did not improve the performance of FIT. When using FIT for triage in primary care, sensitivity is valued over specificity (i.e., reducing false negatives is more valued than reducing false positives). Accordingly, for a model to outperform FIT, additional predictors must enhance the model’s ability to distinguish those with and without cancer specifically within the population who have a FIT value around the FIT threshold. Most people tested will have a negative FIT and many of these individuals will have abnormal blood tests for other reasons. Similarly, many people with a positive FIT will have normal blood tests. For these reasons, although the model fit may improve by including more variables this does not result in better discrimination for colorectal cancer above FIT alone.

### Strengths and Limitations

This study has some strengths. This is the largest cohort of symptomatic primary care patients tested with FIT in the UK and as a result is highly valuable to inform future guidelines around FIT for triage of symptomatic patients. The study comprises tests prior to and during the COVID-19 pandemic and suggests that among primary care tests, the performance of FIT has remained stable throughout. Centralized FIT and blood testing at the the OUHT Clinical Biochemistry Laboratory serves primary care clinicians county-wide and allowed for highly complete assessment of FIT and blood test values. We accessed the referral text to explore the performance of FIT in strata defined by GP reported symptoms.

The prevalence of and type of symptoms reported may have differed if we had accessed primary care records or asked patients to report their symptoms directly^23^. A 6-month follow-up period was used for the primary analysis to optimize the number of cancers included but in sensitivity analyses that aligned with national guideline-setting, 12-month follow-up showed similar results.

With respect to limitations, the sample was restricted to individuals for whom blood test results were available within a 90-day window surrounding FIT. As serum ferritin and CRP results were only available for a subset of those undergoing FIT, we used multiple imputation to compare models generated from stepwise selection in imputed and complete case datasets. Models included FIT as continuous, categorical, and using splines. Blood tests were included as continuous and categorical (abnormal vs. abnormal). In each permutation, different variables were retained suggesting some instability. As no predictive model was identified that performed better than FIT alone, neither internal validation nor optimism correction were pursued. Further work combining large prospective cohorts with uniform collection of common blood tests including CRP and ferritin at baseline could improve our understanding of the predictive importance of these test results.

### Comparison with existing literature

A recent meta-analysis of FIT at a threshold of 10 µg Hb/g faeces estimated a sensitivity of 87.2% and a specificity of 84.4% (n=48,872)^8^. In the current study, the sensitivity and specificity of FIT-alone were 92.1% and 91.5% respectively. The meta-analysis found that the type of reference standard used (colonoscopy or follow-up), the place of recruitment (primary care facility or colonoscopy unit) and CRC prevalence (≥ 3%) were significant sources of heterogeneity in sensitivity and specificity^8^. These factors likely explain, at least in part, the higher sensitivity and specificity observed in this study.

This is the one of two studies to formally and systematically evaluate blood tests in addition to FIT in symptomatic patients, and one of few to analyze FIT supplemented with other variables. The f-Hb, age and sex test score (FAST) was not superior to FIT alone^12^. COLONPREDICT included FIT, rectal bleeding, benign anorectal lesions, rectal mass, serum carcinoembryonic antigen, blood haemoglobin, colonoscopy in the last 10 years, treatment with aspirin, sex and age, and was derived and validated in a higher-risk referred population^24^. The COLONPREDICT model achieved 89% sensitivity and 75.8% specificity, both lower than the performance of FIT alone in this study.

A UK-based study of whether demographic, lifestyle (e.g., smoking, physical activity), or clinical factors (family history, symptoms) could add to the predictive value of FIT found that only family history of polyps showed a significant association once FIT was taken into account^25^. In the current study, family history was not retained in stepwise models, however the indicator was based on the physicians referral notes whereas in the aforementioned study, patients were prospectively asked about family history.

### Implications for research and practice

Adding blood test results to FIT does not appear to improve on the performance of FIT in primary care in a clinically meaningful way. The lack of an apparent age-effect after taking into account FIT suggests that age-specific thresholds for FIT positivity would not improve test performance. Particularly in light of the COVID pandemic and the suspected accumulation of undiagnosed cancers and unscreened adults^11^, effective methods to triage low and/or “intermediate” risk patients to referral are more needed than ever^26^. Our results suggest however, that neither age, nor blood tests, nor clinical indication as recorded by the physician should be used to inform referral to colonoscopy after FIT.

FIT alone is simple, easily implemented and effective to triage patients from primary care to colonoscopy. A key question remains how to detect FIT negative colorectal cancers^8^. Based on the false negatives in this study, no practical rules using blood tests or clinical indication were apparent. For example, while half the false negatives had raised CRP, scoping all patients with raised CRP regardless of FIT would increase the false positives by at least two-fold. Future research may benefit from an agnostic approach to building the prediction model. For example, new predictive markers could be discovered by applying machine learning models to large, representative databases of electronic health records^27^.

Continued research into alternative risk stratification tools to reduce false negatives and false positives is worthwhile. For example, urinary volatile organic compounds may have promise as a second-stage/rule-out test to complement FIT^10 28^. Other risk stratification tools that have been suggested in the screening setting such as polygenic risk scores^29-31^, or emerging technologies such as circulating and/or faecal tumor DNA^32^, could also be explored in combination with FIT for triage of primary care patients.

In the interim, follow-up care of FIT-negative patients should likely focus on safety netting, including re-evaluation of persistent and additional symptoms within a pre-specified timeframe, and potentially re-testing with FIT after a set time interval has passed, as data is lacking to support immediate repeat FIT testing at this time^33-35^.

## Supporting information

Ethics

## Data Availability

Data were retrieved from Oxford University Hospitals NHS Foundation Trust (OUH) and are not publicly available.

## Author contributions

Diana R Withrow: conceptualisation, data curation, formal analysis, investigation, methodology, visualisation, writing – original draft, and writing – review & editing

Brian Shine: data curation, methodology, resources, writing – review & editing

Jason Oke: conceptualisation, data curation, formal analysis, funding acquisition, investigation, methodology, supervision, and writing – review & editing

Andres Tamm: formal analysis, methodology, visualisation, writing – review & editing

Tim James: data curation, methodology, project administration, writing – review & editing

Eva Morris: supervision, writing – review & editing

Jim Davies: data curation, writing – review & editing

Steve Harris: data curation, writing – review & editing

James East: conceptualisation, funding acquisition, supervision, writing – review & editing

Brian D Nicholson: conceptualisation, formal analysis, funding acquisition, investigation, methodology, project administration, supervision, writing – original draft, and writing – review & editing

## Ethics committee approval

This study was conducted as a service evaluation with registration, review and approval process within the OUH Datix governance structure (Service evaluation registration identifier: CSS-BIO-3-4730). As service evaluation, this work is not subject to the Department of Health’s UK Policy Framework for Health and Social Care Research (2017). The Sponsorship and Ethics Lead within the Research Governance, Ethics and Assurance Team in the Research Support Office at the University of Oxford has confirmed that it requires neither sponsorship nor research ethics review.

## Acknowledgements

This research uses data provided by patients and collected by the NHS as part of their care and support. We appreciate feedback from Patrick McGuire. The views expressed are those of the authors and not necessarily those of the National Health Service, the NIHR or the Department of Health.

