## Supplementary figures and images for "Combining faecal immunochemical testing with blood test results to identify patients with symptoms at risk of colorectal cancer: a consecutive cohort of 16,604 patients tested in primary care"

### Supplementary materials

**SUPPLEMENTARY TABLES**

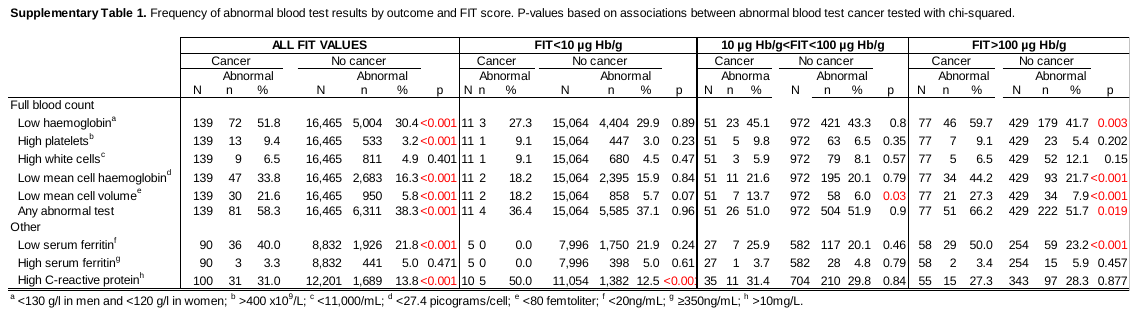

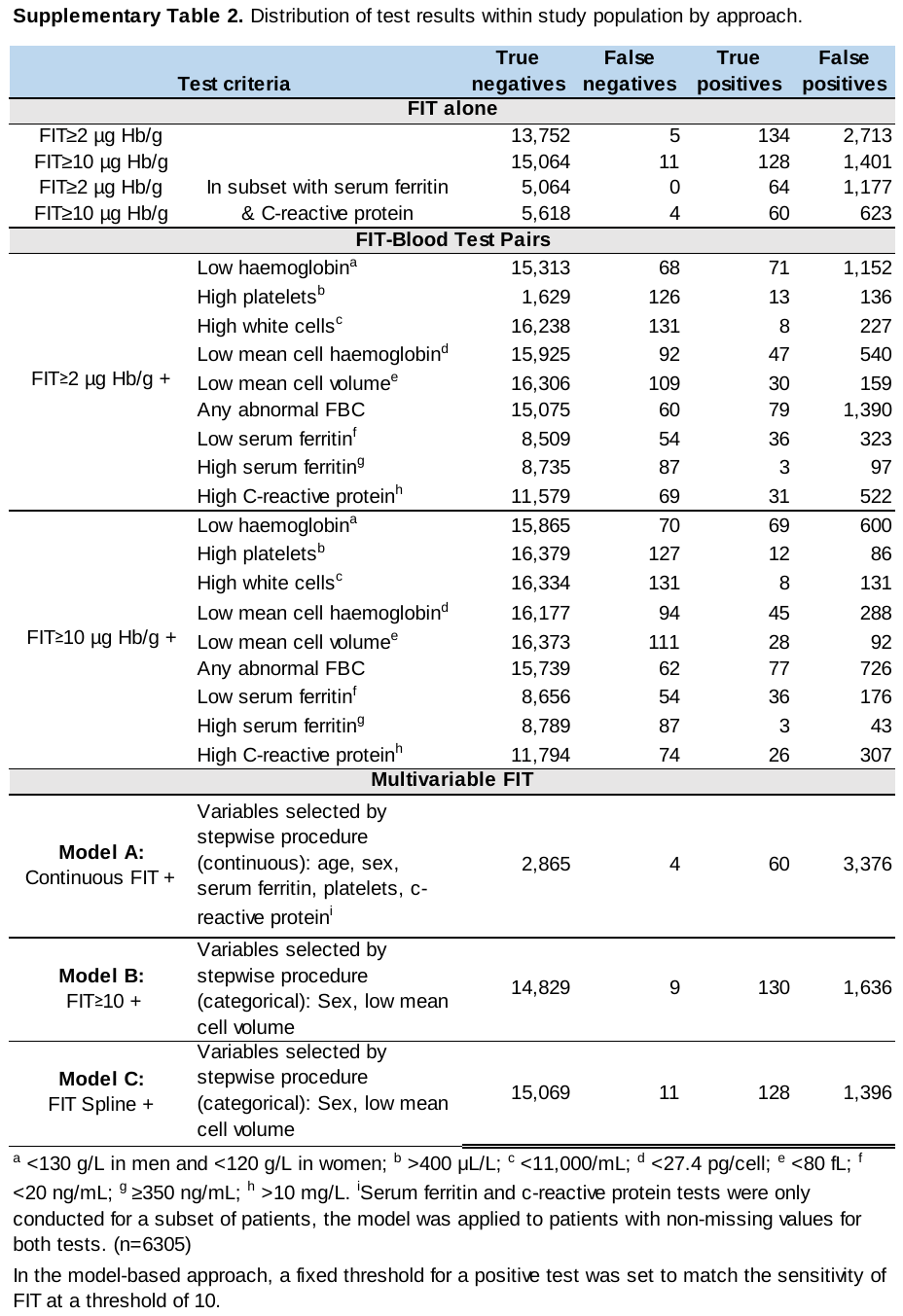

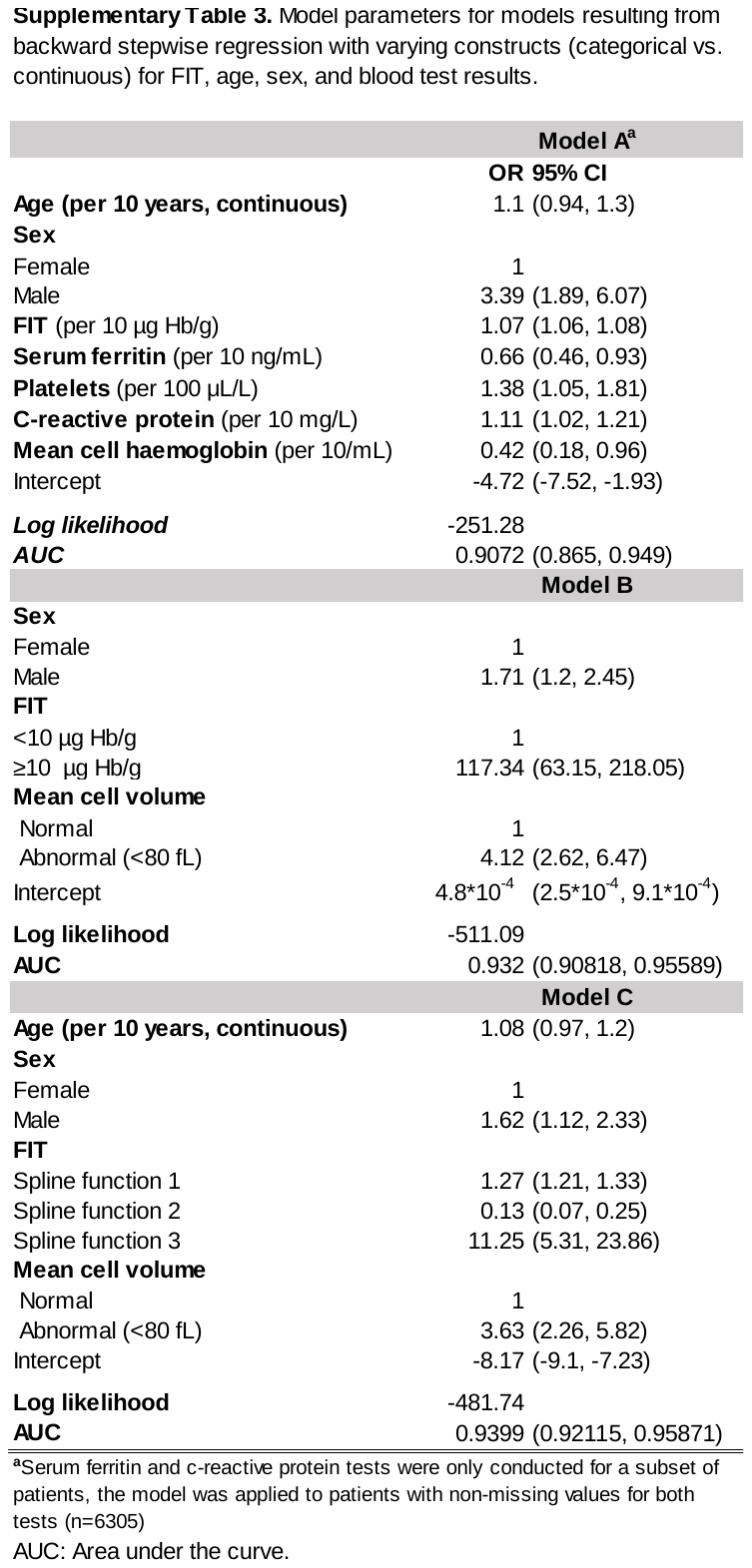

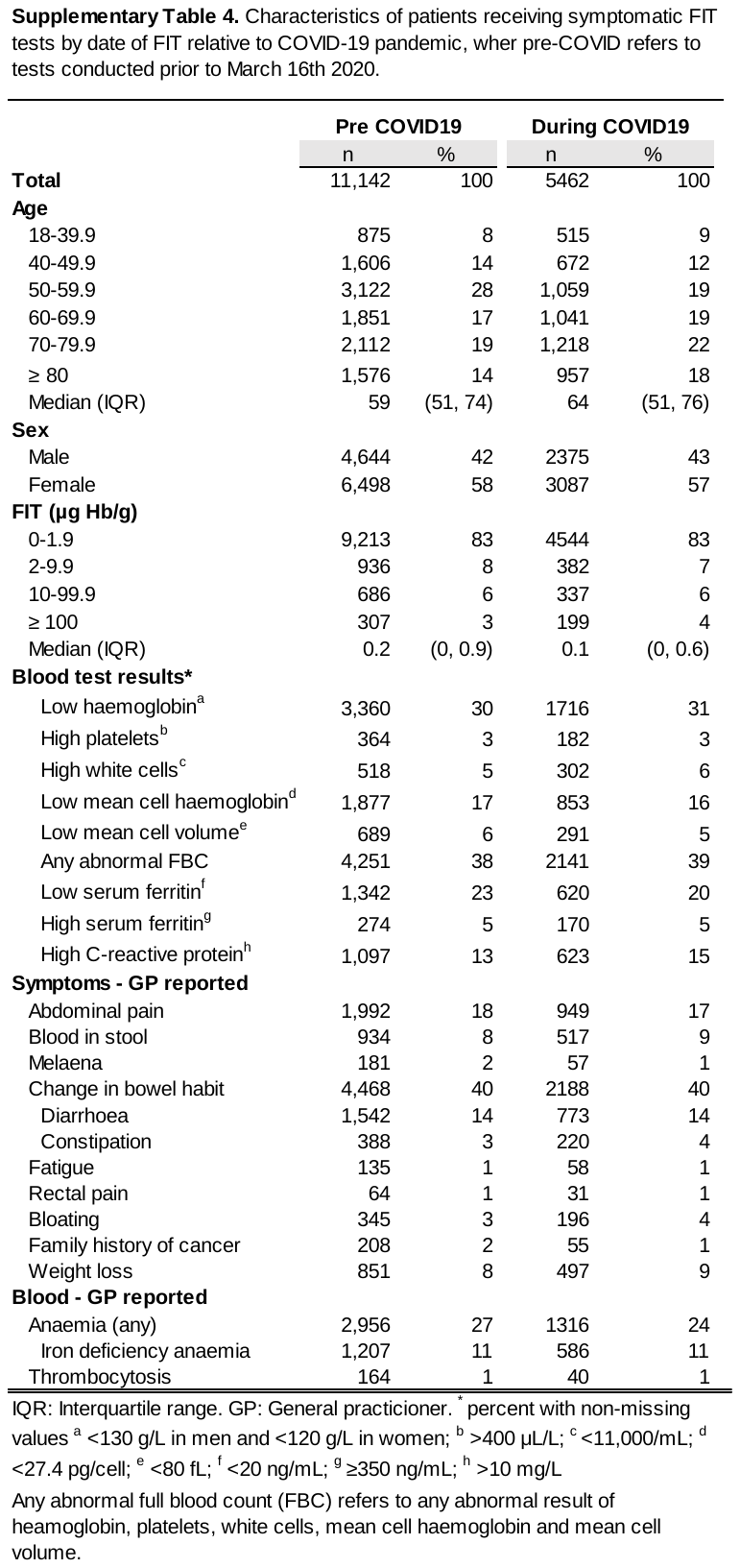

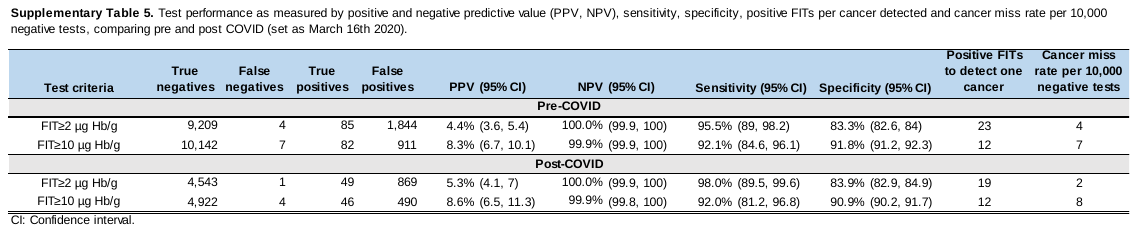

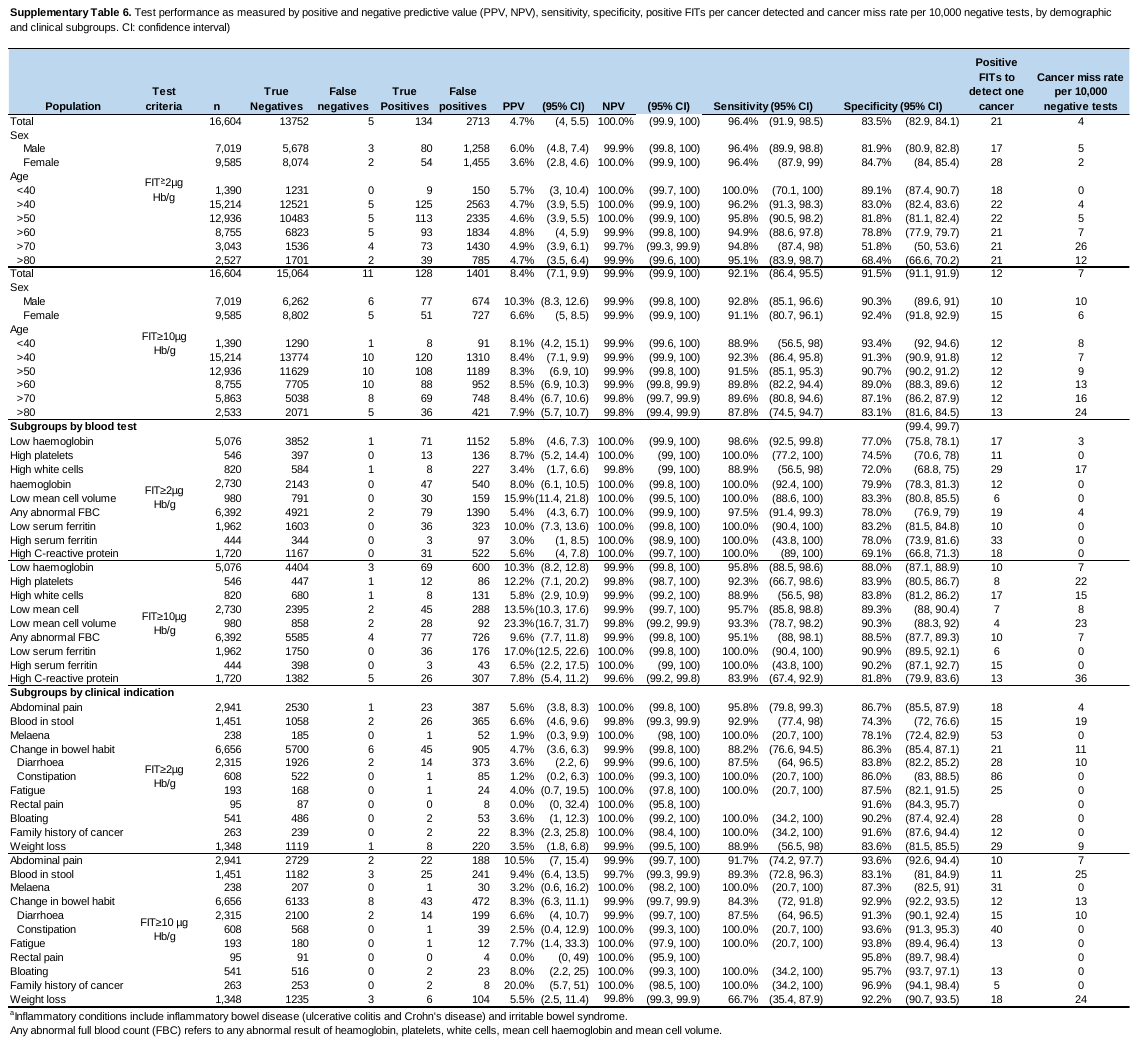

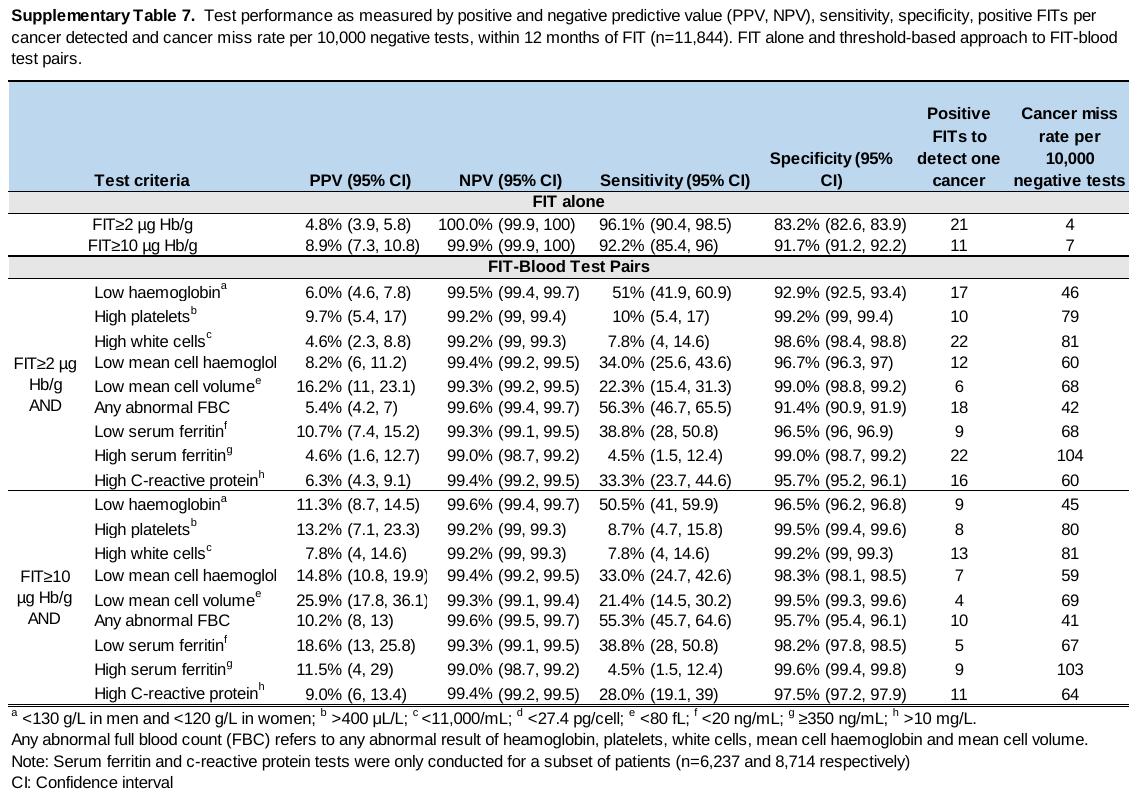

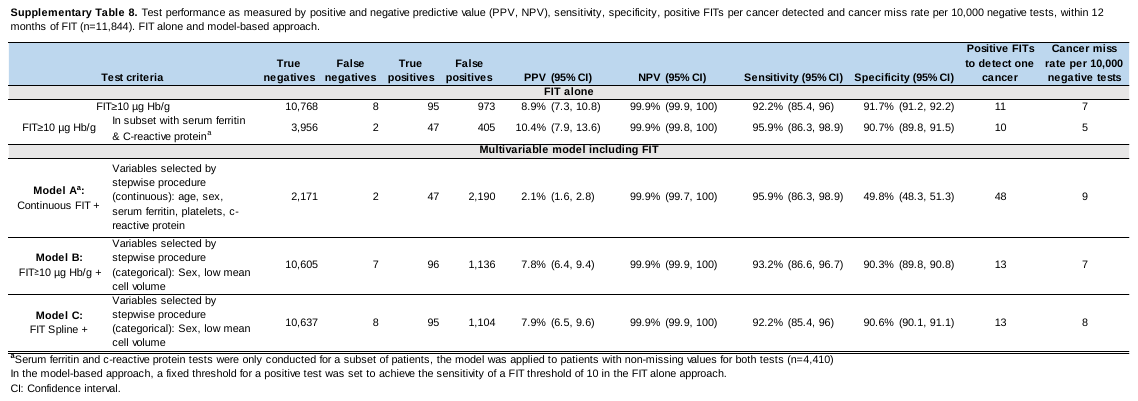
