## Supplementary material for "Combining faecal immunochemical testing with blood test results to identify patients with symptoms at risk of colorectal cancer: a consecutive cohort of 16,604 patients tested in primary care": Ethics

UNIVERSITY OF OXFORD

Joint Research Office
Boundary Brook House
Churchill Drive, Headington, Oxford OX3 7GB

09 October 2021

To whom it may concern:

**Title: Combining faecal Immunochemical testing with blood test results to identify patients with symptoms at risk of colon cancer: a consecutive cohort of 15,504 patients tested in primary care**

This letter is to confirm that the work referenced above was conducted as a service evaluation with registration, review and approval process within the OUH Datix governance structure (Service evaluation registration identifier: CSS-BIO-3-4730).

As service evaluation, this work is not subject to the Department of Health's *UK Policy Framework for Health and Social Care Research* (2017). It requires neither sponsorship nor research ethics review.

This opinion can be reviewed by reference to the HRA's algorithm, available at http://www.hra-decisiontools.org.uk/research/ and attendant leaflet, *Defining Research,* or by reference to The Health Care Quality Improvement Partnership (HQIP)'s *Guide for Clinical Audit, Research and Service review.*

Should you require further information, please do not hesitate to contact me.

Sincerely,

Karen Melham

Dr Karen Melham
Sponsorship and Ethics Lead
Research Services
University of Oxford

Copy to: Ms Jo Franklin, Research Governance Manager, OUH NHS Foundation Trust

https://researchsupport.admin.ox.ac.uk/ctrg
